# Efficacy and safety of praziquantel plus Artemisinin-based combinations in the treatment of Kenyan children with *Schistosoma mansoni* infection: an open-label, randomized, head-to-head, non-inferiority (SCHISTOACT) trial

**DOI:** 10.1101/2024.01.12.24301192

**Authors:** Charles O. Obonyo, Vincent O. Were, Peter Wamae, Erick M.O. Muok

## Abstract

**Background:** Praziquantel alone is insufficient for the control of schistosomiasis. Unlike praziquantel, artemisinin derivatives are effective for treating juvenile schistosome worms but not adult worms. Few studies have assessed the role of combination therapy, including praziquantel and artemisinin-based combinations, in treating schistosomiasis.

**Methods:** A randomized, open-label, noninferiority trial was conducted in central Kenya to assess the efficacy and safety of praziquantel plus one of four artemisinin-based combination therapies in treating intestinal schistosomiasis. 540 children aged 9-15 years with *Schistosoma mansoni* infection were randomly assigned (1:1:1:1:1) to receive a single oral dose of praziquantel (40mg/kg/day) alone or in combination with a 3-day course of artesunate plus sulfalene/pyrimethamine or artesunate plus amodiaquine or artesunate plus mefloquine or dihydroartemisinin-piperaquine. The primary endpoint was the cure rate assessed at six weeks in a per-protocol population. The noninferiority margin was defined as the lower limit of 95%CI of the risk difference in cure rates less than -10%.

**Results:** Cure rates were available for 523 children. Overall, 82.5%, 81.7%, 76.2%, 88.7% and 85.7% of patients on praziquantel, praziquantel-artesunate-sulfalene/pyrimethamine, praziquantel-artesunate-amodiaquine, praziquantel-artesunate-mefloquine, and praziquantel-dihydroartemisinin-piperaquine, respectively, were cured. Non-inferiority was declared for praziquantel-artesunate-mefloquine (difference 6.2 [95%CI -3.3 to 15.6]) and praziquantel-dihydroartemisinin-piperaquine (3.2 [-6.7 to 13.1]) but not for praziquantel-artesunate-sulfalene/pyrimethamine (-0.8 [-11.2 to 9.6]) or praziquantel-artesunate-amodiaquine (-6.3 [-17.3 to 4.6]). A significantly lower number of adverse events were reported in the praziquantel arm than in the combined treatment arm. No serious adverse events were observed.

**Conclusions:** Praziquantel-dihydroartemisinin-piperaquine and praziquantel-artesunate-mefloquine are suitable alternatives to praziquantel monotherapy. The role of artemisinin-based combinations in the treatment of intestinal schistosomiasis remains unclear.

**Clinical trials registration:** Pan-African Clinical Trials Registry, PACTR202001919442161.

## INTRODUCTION

Schistosomiasis, a disease caused by trematode worms of the genus *Schistosoma,* is a major global health issue. Over 90% of the 240 million people infected reside in sub-Saharan Africa, where S*. mansoni* and *S. haematobium* are prevalent [1]. The greatest burden of schistosomiasis is in school-age children, with consequences that include anaemia, school absenteeism, impaired child growth, physical fitness, and cognitive and intellectual development. The global schistosomiasis control strategy relies on praziquantel as it is effective against all schistosome species that infect human populations, is administered as a single oral dose, is affordable, and is safe [2]. Praziquantel is effective against adult schistosome worms but ineffective against the parasite’s juvenile stages and does not prevent reinfection [3]. Reliance on a single drug for wide-scale chemotherapy poses a risk of the emergence of praziquantel-resistant parasites.

The drug development pipeline for schistosomiasis is empty, highlighting the urgent need for new treatments in addition to praziquantel. Artemisinins, which are highly effective against malaria, have shown specific activity against juvenile worms of schistosomiasis [4,5]. While artemisinin-based combination treatments (ACTs) are less effective than praziquantel alone, combining praziquantel with an artemisinin derivative improved efficacy in treating schistosomiasis [6]. These differences in drug action provide a basis for combining praziquantel with ACTs to enhance praziquantel efficacy and prevent drug resistance development.

At least five ACTs have been approved for treating uncomplicated malaria [7], but their comparative efficacy against schistosomiasis, combined with praziquantel, has been evaluated in only two studies with mixed results [8, 9]. The SCHISTOACT study evaluated the effectiveness of different treatment options for schistosomiasis in combination with praziquantel. The study assessed the efficacy and safety of praziquantel alone or in combination with one of four ACT combinations, namely artesunate plus amodiaquine (As+AQ), artesunate plus sulfalene-pyrimethamine (As+SP), artesunate plus mefloquine (As+MQ), or dihydroartemisinin-piperaquine (DHAP) for treating *S. mansoni* in Kenyan children. The four ACTs were selected due to their wide availability, co-formulation, ability to be dosed once daily, well-established safety profile, and their previous evaluation for the treatment of schistosomiasis with inconclusive results [8–14]. The control group received only praziquantel.

## PATIENTS AND MATERIALS

### Study design and population

The SCHISTOACT study is a phase III, open-label, head-to-head, randomized controlled, noninferiority trial conducted in seven primary schools within the Mwea sub-count**y** in Kirinyaga County, central Kenya. The study area is endemic to *S. mansoni* and has extremely low malaria transmission (<1/1000 individuals) [15].

### Eligibility criteria

This study enrolled children aged 9 to 15 years who had tested positive for *S. mansoni* infections and were willing to participate in follow-up evaluations. We excluded children weighing over 50 kg, those co-infected with *P. falciparum*, those with severe malnutrition (severe wasting or mid-upper arm circumference <12cm), those with severe anaemia (haemoglobin level < 8.0 g/dL) or those with a severe illness (e.g., epilepsy). Additionally, children with a known hypersensitivity to artesunates, sulfonamides, mefloquine, or praziquantel were excluded. Patients who had received an antimalarial or anti-schistosomal drug within 28 days before enrollment in the study were also excluded.

### Sample size calculation

We hypothesized that combination therapy was not inferior to PZQ in treating schistosomiasis. We assumed a cure rate of 72% with praziquantel from longitudinal studies in western Kenya [16]. The noninferiority margin was set at -10 percentage points for the risk difference in cure rates, which was considered clinically equivalent. With this noninferiority margin, a power of 80%, and a one-sided alpha value of 0.025, a total sample size of 540 (108 in each study arm) was needed after accounting for a dropout rate of 5%.

### Laboratory procedures

Stool samples from children at enrolment, week six, and week 12 were assessed. Duplicate slides from one stool sample were prepared and examined independently under the microscope by two skilled laboratory technicians. The *S. mansoni* egg and soil-transmitted helminth egg were quantified with the Kato-Katz faecal smear technique. A template containing approximately 41.7 mg faeces was filled, the number of *S. mansoni* eggs per slide was counted, and the mean of the two slides was multiplied by 24 and expressed as eggs per gram of faeces (EPG). The infection intensity was classified as light (1-99 epg), medium (100-399 epg) or heavy (>400 epg) [17]. A third technician read 10% of the slides and all slides, where the initial readings varied between the two technicians by more than 20%. A capillary blood sample was taken from the fingerprick for the estimation of haemoglobin (HemoCue) and malaria parasites (by microscopy). Children whose stools tested positive for *S. mansoni* and who met all the eligibility criteria were enrolled in the study.

### Treatment and follow-up

A computer-generated randomization list was used to randomly assign children into five treatment groups (1:1:1:1:1), divided into blocks of 15. The study drugs were pre-packaged in opaque, sealed envelopes and labelled with the randomization/study number per the treatment assigned. A medical history and clinical examination were conducted by the study clinician, which included weight and height measurements. All enrolled children were assigned a study number, and the study nurse administered the treatment to each child by opening the sealed envelope. The laboratory technicians were masked to the treatment assignment. The patients and clinicians were not masked to the intervention groups.

All the study drugs were purchased from a local chemist in Kisumu. At enrolment, each child was provided with a single dose PZQ (Biltricide, Bayer Healthcare, Leverkusen, Germany) tablet containing 40mg/kg of PZQ to the nearest quarter tablet (600mg tablets). Additionally, children assigned to the (i) PZQ plus As+SP group, received As+SP (Coarinate Junior FDC, Dafra Pharma, Turnhout, Belgium) tablets containing 4mg/kg/day of the artesunate component once daily for three days. As+SP is a fixed-dose combination tablet consisting of 100mg artesunate, 250mg sulfalene, plus 12.5mg pyrimethamine (pack of three tablets). (ii) The PZQ plus As+AQ group, received As+AQ (ASAQ, Winthrop, Sanofi-Aventis, France) tablets containing 4mg/kg/day of the artesunate component once daily for three days. As+AQ is a fixed-dose combination tablet consisting of 100mg of artesunate and 270mg of amodiaquine. (iii) The PZQ plus As+MQ group, received As+MQ (Artequin, Acino/Mepha, Switzerland) tablets containing 4mg/kg/day of the artesunate and 8.3mg/kg/day of mefloquine once daily for three days. As+MQ is a fixed-dose combination tablet consisting of 200 mg artesunate and 250mg mefloquine. (iv) The PZQ plus DHAP group, was administered DHAP (D-ARTEPP, Guilin Pharmaceutical Co. Ltd, China) tablets containing 4mg/kg/day of dihydroartemisinin and 20mg/kg/day of piperaquine once daily for three days. DHAP is a fixed-dose combination tablet containing 40mg of dihydroartemisinin and 320mg of piperaquine.

Before drug administration, all children were provided food to reduce the nauseating effect of the study drugs and improve their bioavailability [18]. The study nurse administered all the study drugs orally. Children were observed for one-hour post-treatment for acute adverse events. If vomiting occurred within one hour of drug ingestion, a repeat dose was given. According to the national treatment guidelines, all children with soil-transmitted infections received a single dose of 400mg of albendazole.

Children were followed up at six and 12 weeks after starting treatment. Stool samples were collected at six and 12 weeks to check for *S.mansoni* infection. The primary outcomes were assessed six weeks post-treatment to correspond with the time of schistosome larval maturation. By the end of the study, all children received a single dose of 40mg/kg PZQ.

### Study outcomes

The primary endpoint was the cure rate, which measured the proportion of children in each treatment arm who were not shedding eggs at week six post-treatment. Secondary endpoints included the percentage of participants who excreted eggs, the cumulative cure rate at week 12, the intensity of infection, the egg reduction rate, reinfection, and adverse events within 24 hours post-treatment. The arithmetic mean egg count (AM) was calculated from the baseline and post-treatment data. The reinfection rate was defined as the proportion of children cured at week 6 who had new infections at week 12. The egg reduction rate was defined as the percentage reduction in the mean number of S. mansoni eggs between baseline and post-treatment and was calculated as 1-[AM egg count post-treatment/AM egg count at enrollment] x100. Adverse events were assessed by the clinician one hour after ingestion of the study treatment and 24 hours later. The adverse events were classified as mild, moderate, or severe based on their intensity and their effect on daily activities.

### Statistical analysis

We analyzed data using IBM SPSS for Windows version 20.0 and STATA version 12.0 packages. Baseline characteristics and outcomes data were analyzed for all children who received at least one dose of the study drug, whether they completed the study or not.

Pearson’s Χ^2^ test was used to compare cure rates (CRs) and adverse events between different treatment arms. The results were summarized as relative risks (RRs) or risk differences with 95% confidence intervals (CIs). Noninferiority was determined if the lower bound of the 95%CI for the risk difference did not exceed -10. The primary outcome was analyzed using both the per-protocol and intention-to-treat approaches, with per-protocol analysis being the main approach.

Changes in continuous variables were assessed using a one-way analysis of variance, and post-hoc pairwise comparisons between group means were performed using the least significant difference. The frequency and pattern of adverse events were examined in the intent-to-treat population. Statistical significance was defined as two-sided p-values less than 0.05.

## RESULTS

A total of 2003 children were screened for *S. mansoni* infection between September 2018 and November 2018. 806 (40.2%) children tested positive for *S. mansoni,* and none tested positive for malaria. 540 children were enrolled and randomized into five study groups (Figure 1). After randomization, 17 (3.1%) children were lost to follow-up, and another 85 (15.7%) were lost to follow-up between weeks six and 12.

**Figure 1.**
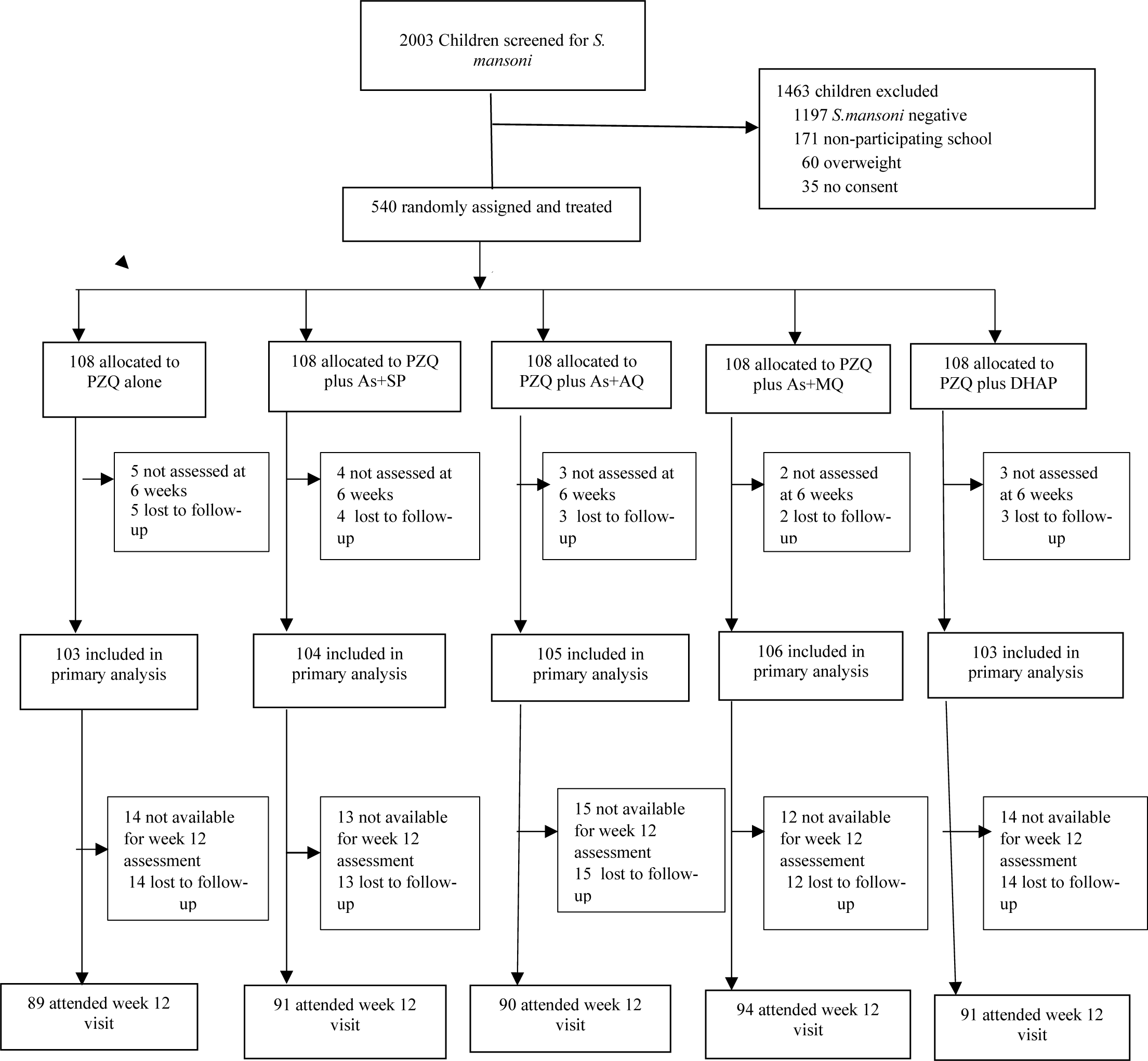
The study profile. PZQ=praziquantel; As+SP=artesunate plus sulfalene/pyrimethamine; As+AQ= artesunate plus amodiaquine; As+MQ=artesunate plus mefloquine; DHAP=Dihydroartemisinin-piperaquine

The mean age of enrolled children was 10.6 (SD 1.4) years, and 47.8% (258/540) were female. Overall, the intensity of infection was light 344 (63.7%), moderate 140 (25.6%) or heavy 54 (10.4%). Treatment arms were balanced regarding age, sex, body weight, and baseline infection intensity. Table 1 is a summary of the baseline characteristics.

**Table 1.** Baseline characteristics of the study participants. Key: N=number; CI=confidence interval; SD=standard deviation; PZQ=praziquantel; As+SP=artesunate plus sulfalene/pyrimethamine; As+AQ= artesunate plus amodiaquine; As+MQ=artesunate plus mefloquine; DHAP=Dihydroartemisinin-piperaquine

| Variable | PZQ alone | PZQ plus As+SP | PZQ plus<br>As+AQ | PZQ plus As+MQ | PZQ plus DHAP |
| --- | --- | --- | --- | --- | --- |
| **Number** | 108 | 108 | 108 | 108 | 108 |
| **Sex** |  |  |  |  |  |
| Male, N (%) | 54 (50.0) | 61 (56.5) | 60 (55.6) | 54 (50) | 53 (49.1) |
| Female, N (%) | 54 (50.0) | 47 (43.5) | 48 (44.4) | 54 (50) | 55 (50.9) |
| **Age (years)** |  |  |  |  |  |
| Mean (SD) | 10.6 (1.3) | 10.6 (1.4) | 10.6(1.4) | 10.6(1.4) | 10.5 (1.4) |
| **Age group** |  |  |  |  |  |
| <12 years, N (%) | 82 (75.9) | 76 (70.4) | 76 (70.4) | 77 (71.3) | 83 (76.9) |
| ≥ 12years, N (%) | 26 (24.1) | 32 (29.6) | 32 (29.6) | 31 (28.7) | 25 (23.1) |
| **Primary schools** |  |  |  |  |  |
| Thiba, N (%) | 14 (13.0) | 16 (14.8) | 13(12.0) | 13 (12.0) | 15 (13.9) |
| Mbui-Njeru, N (%) | 16 (14.8) | 17 (15.7) | 13(12.0) | 16 (14.8) | 17 (15.7) |
| Mukou, N (%) | 19 (17.6) | 18 (16.7) | 20(18.5) | 21(19.4) | 20 (18.5) |
| Mianya, N (%) | 20 (18.5) | 18 (16.7) | 19(17.8) | 18(16.7) | 17 (15.7) |
| Nyangati, N (%) | 10 (9.3) | 9 (18.3) | 11(10.2) | 10 (9.3) | 10 (9.3) |
| Murubara, N (%) | 18 (16.7) | 18 (16.7) | 20 (18.5) | 19 (17.6) | 18 (16.7) |
| Kamuchege, N (%) | 11 (10.2) | 12 (11.1) | 12 (11.1) | 11 (10.2) | 11 (10.2) |
| **Class** |  |  |  |  |  |
| 3, N (%) | 32(29.6) | 32 (29.6) | 30 (27.8) | 29(26.9) | 30(27.8) |
| 4, N (%) | 38(35.2) | 38(35.2) | 33 (30.6) | 41 (38.0) | 36 (33.3) |
| 5, N (%) | 38(35.2) | 38(35.2) | 45 (41.7) | 38 (35.2) | 42 (38.9) |
| **Body weight (Kg)** |  |  |  |  |  |
| Mean (SD) | 29.5 (5.5) | 29.2 (4.7) | 30.6 (5.8) | 30.6 (6.1) | 29.9 (5.9) |
| **Haemoglobin (g/dL)** |  |  |  |  |  |
| Mean (SD) | 13.4 (1.1) | 13.2 (1.1) | 13.3 (1.1) | 13.2 (1.2) | 13.2 (1.1) |
| **Infection intensity** |  |  |  |  |  |
| Light, N (%) | 59 (54.5) | 70 (64.8) | 72 (66.7) | 67(62.0) | 76 (70.4) |
| Moderate, N (%) | 30 (27.8) | 27 (25.0) | 25 (23.1) | 34(31.5) | 24(22.2) |
| Heavy, N (%) | 19 (17.6) | 11 (10.2) | 11 (10.2) | 7 (6.5) | 8 (7.4) |
| **Egg count on day 0** |  |  |  |  |  |
| Arithmetic mean egg count<br>(epg, 95%CI) | 255.2<br>(166.8 to 343.4) | 294.9<br>(122.9 to 466.8) | 204.2<br>(104.8 to 303.6) | 162.7<br>(99.2 to 226.1) | 136.9<br>(81.7 to 192.1) |
| Median | 72 | 48 | 60 | 60 | 48 |

523 (96.9%) of the 540 children included were analyzed for the primary outcome, and 434 (80.4%) were cured. A cure was attained in 82.5%, 81.7%, 76.2%, 88.7%, and 85.7% of children who received PZQ alone, PZQ plus As+SP, PZQ plus As+AQ, PZQ plus As+MQ and PZQ plus DHAP, respectively. Noninferiority was declared for PZQ plus As+MQ (difference 6.2 [95%CI - 3.3 to 15.6]) and PZQ plus DHAP (3.2 [-6.7 to 13.1]) but not for PZQ plus As+SP or PZQ plus As+AQ. The findings of the primary analysis were confirmed by an intention-to-treat analysis (Table 2).

**Table 2.** Summary of Primary Efficacy Endpoints. Key: PZQ=praziquantel; As+SP=artesunate plus sulfalene/pyrimethamine; As+AQ= artesunate plus amodiaquine; As+MQ=artesunate plus mefloquine; DHAP=Dihydroartemisinin-piperaquine; n= number; N=total number; CI=confidence interval; ITT=intention-to-treat; PP=per-protocol; Ref=reference.

| Variable | PZQ alone | PZQ plus AS+SP | PZQ plus As+AQ | PZQ plus As+MQ | PZQ plus DHAP |
| --- | --- | --- | --- | --- | --- |
| **Cure rate on week 6 [PP],**<br>n/N (%) [95%CI] | 85/103 (82.5%)<br>[75.2 to 89.5%] | 85/104 (81.7%)<br>[74.3 to 89.1%] | 80/105 (76.2%)<br>[68.1 to 84.3%] | 94/106 (88.7%)<br>[82.7 to 94.7%] | 90/105 (85.7%)<br>[79.0 to 92.4%] |
| Risk ratio (95%CI) | Ref | 0.99 (0.87 to 1.12) | 0.92 (0.80 to 1.06) | 1.07 (0.96 to 1.20) | 1.04 (0.92 to 1.17) |
| Risk difference (95%CI) | Ref | -0.80<br>(-11.2 to 9.6) | -6.33<br>(-17.3 to 4.6) | 6.15<br>(-3.3 to 15.6) | 3.19<br>(-6.74 to 13.1) |
| **Cure rate on week 6 [ITT],**<br>n/N (%) [95%CI] | 85/108 (78.7%)<br>[69.9 to 85.5%] | 85/108 (78.7%)<br>[69.9 to 85.5%] | 80/108 (74.1%)<br>[64.9 to 81.5%] | 94/108 (87.0%)<br>[79.2 to 92.2%] | 90/108 (83.3%)<br>[75.0 to 89.3%] |
| Risk ratio (95%CI) | Ref | 1.0 (0.87 to 1.14) | 0.94 (0.81 to 1.09) | 1.11 (0.98 to 1.25) | 1.06 (0.93 to 1.21) |
| Risk difference (95%CI) | Ref | 0<br>(-10.9 to 10.9) | - 4.6<br>(-15.9 to 6.68) | 8.3<br>(-1.65 to 18.3) | 4.6<br>(-5.8 to 15.1) |
| **Cure rate by baseline infection intensity** |  |  |  |  |  |
| Light | 50/56 (89.3) | 61/67 (91.0) | 60/69 (87.0) | 59/66 (89.4) | 66/73 (90.4) |
| Moderate | 24/29 (82.8) | 18/26 (69.2) | 15/25 (60.0) | 28/33 (84.8) | 20/24 (83.3) |
| Heavy | 11/18 (61.1) | 6/11 (54.5) | 5/11 (45.5) | 7/7 (100) | 4/8 (50) |

The ERR at six weeks was 84.2% with PZQ alone but above 90% in the four combination treatment arms (Table 3). Mean differences in egg count were increased when PZQ was combined with either As+MQ or DHAP but not when combined with As+SP or As+AQ (Table 3). In all the treatment groups, the intensity of infection decreased significantly at week six post-treatment (Figure 2, panel B).

**Table 3.** Summary of secondary endpoints. PZQ=praziquantel; As+SP=artesunate plus sulfalene/pyrimethamine; As+AQ= artesunate plus amodiaquine; As+MQ=artesunate plus mefloquine; DHAP=Dihydroartemisinin-piperaquine; n= number; N=total number; CI=confidence interval; Δ = mean difference; EPG= eggs per gram; Ref=reference;1=egg reduction rate at 6 weeks; 2= egg reduction rate at 12 weeks

|  | PZQ alone | PZQ plus AS+SP | PZQ plus As+AQ | PZQ plus As+MQ | PZQ plus DHAP |
| --- | --- | --- | --- | --- | --- |
| **Cure rate on week 12,**<br>n/N (%) [95%CI] | 59/89 (66.3)<br>[55.8% to 75.4%] | 57/91 (62.6)<br>[52.2% to 72.0%] | 52/90 (57.8)<br>[45.1% to 65.1%] | 52/94 (55.3)<br>[39.2% to 59.7%] | 45/91 (49.5)<br>[47.3% to 67.6%] |
| Risk ratio (95%CI) | Ref | 0.94 (0.76 to 1.17) | 0.87 (0.69 to 1.09) | 0.83 (0.66 to 1.05) | 0.75 (0.58 to 0.96) |
| **Arithmetic mean EPG** |  |  |  |  |  |
| Baseline EPG (95%CI) | 255.2<br>(166.8 to 343.4) | 294.9<br>(122.9 to 466.8) | 204.2<br>(104.8 to 303.6) | 162.7<br>(99.2 to 226.1) | 136.9<br>(81.7 to 192.1) |
| Mean difference in egg<br>count at 6 weeks, Δ, (95%CI),<br>p-value | Ref | 25.8 (-3.7 to 55.3),<br>0.09 | 24.5 (-4.9 to 53.9),<br>0.10 | 35.5 (6.2 to 64.9),<br>0.02 | 35.7 (6.3 to 65.1),<br>0.02 |
| Mean EPG at six weeks<br>(95%CI) | 40.3<br>(1.19 to 81.7) | 14.5<br>(1.83 to 27.2) | 15.8<br>(2.14 to 33.7) | 4.73<br>(0.43 to 9.91) | 4.57<br>(0.64 to 8.49) |
| Egg reduction rate (%)<sup>1</sup> | 84.2<br>(77.2 to 91.2) | 95.1<br>(90.9 to 99.2) | 92.2<br>(87.1 to 97.3) | 97.1<br>(93.9 to 100) | 96.7<br>(93.2 to 100) |
| Mean difference in egg<br>count at 12 weeks, Δ,<br>(95%CI), p-value | Ref | 104.1 (-5.1 to<br>213.2), 0.06 | 62.1 (-47.4 to<br>171.5), 0.27 | 82.6 (-25.8 to 190.9),<br>0.14 | 111.0 (1.87 to<br>220.2), 0.046 |
| Mean EPG at 12 weeks<br>(95%CI) | 234.3<br>(130.9 to 337.8) | 130.29<br>(64.7 to 195.9) | 172.27<br>(98.4 to 246.1) | 151.78<br>(70.6 to 232.9) | 123.29<br>(66.6 to 179.9) |
| Egg reduction rate (95%CI)<sup>2</sup> | 8.2 (2.50 to 13.9) | 55.8 (45.6 to 66.0) | 15.6 (8.10 to 23.1) | 6.7 (1.65 to 11.8) | 9.9 (3.16 to 16.0) |
| **Re-infection rate, n/N (%)** | 28/89 (31.5) | 31/91 (34.1) | 30/90 (33.3) | 39/94 (41.5) | 43/91 (47.3) |
| Risk ratio (95%CI) | Ref | 1.08 (0.71 to 1.65) | 1.06 (0.63 to 1.62) | 1.32 (0.89 to 1.95) | 1.50 (1.03 to 2.19) |
| **Incidence of adverse events** |  |  |  |  |  |
| One adverse event | 13 | 22 | 13 | 20 | 17 |
| 2+ adverse events | 2 | 13 | 16 | 11 | 11 |
| Number of children with<br>adverse events, N (%) | 15 (13.9) | 35 (32.4) | 29 (26.9) | 31 (28.7) | 28 (25.9) |
| Risk ratio (95%CI) | Ref | 2.33 (1.39 to 3.91) | 1.93 (1.12 to 3.34) | 2.07 (1.21 to 3.53) | 1.87 (1.07 to 3.24) |

**Figure 2.**
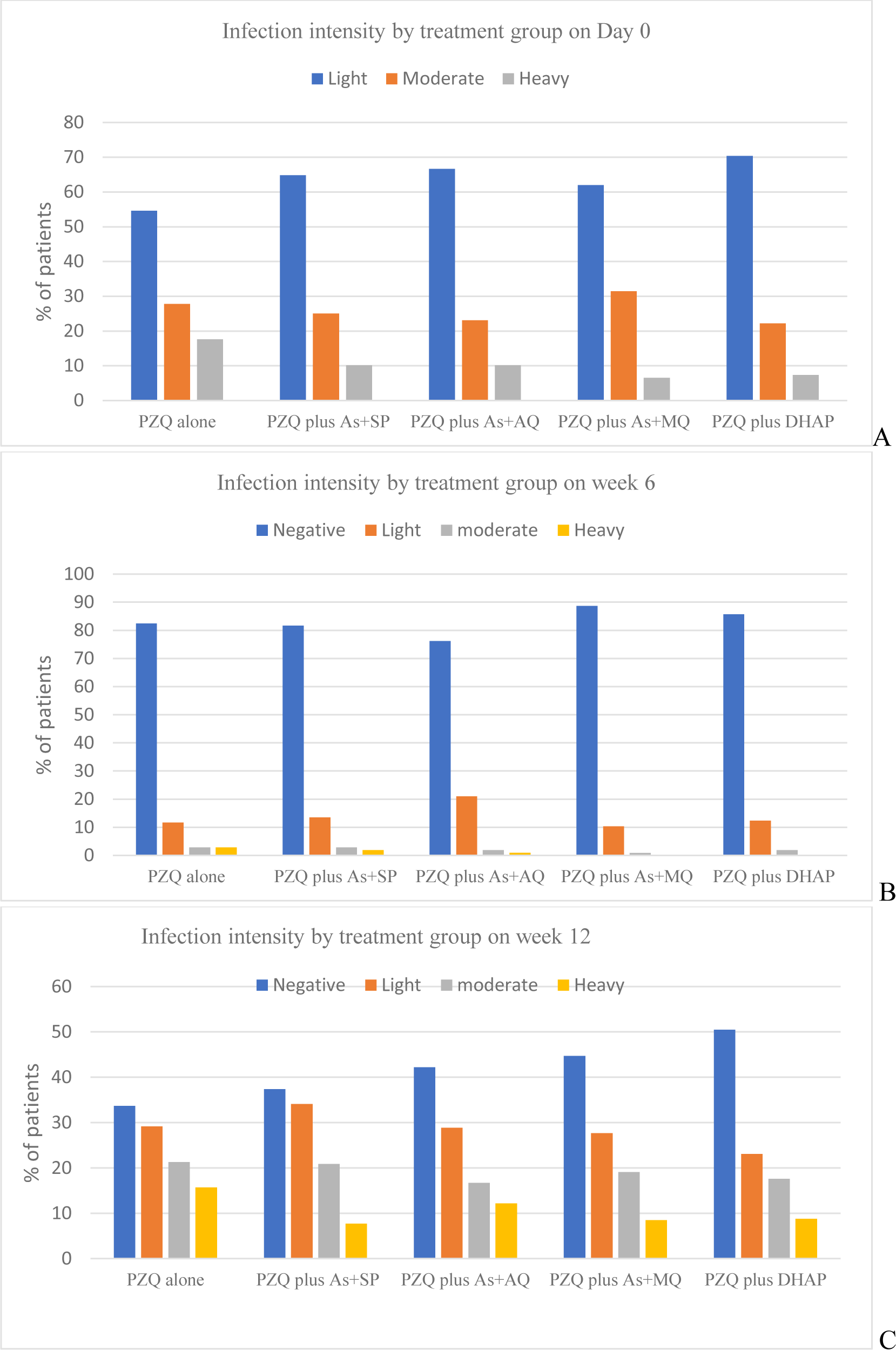
Changes in infection intensity at baseline, six and 12 weeks. PZQ=praziquantel; As+SP=artesunate plus sulfalene/pyrimethamine; As+AQ= artesunate plus amodiaquine; As+MQ=artesunate plus mefloquine; DHAP=Dihydroartemisinin-piperaquine; A=At baseline; B=at 6 weeks follow up; C=at 12 weeks after treatment

At week 12, 455 (84.3%) children were analyzed, and 265 (58.2%) were cured. The cumulative cure rates were much lower than at week six but were comparable between the treatment groups, except for the PZQ plus DHAP group, which was significantly lower than the PZQ alone (RR=0.75, 95%CI 0.58 to 0.96). See Table 3.

190 (43.4%) children were excreting schistosome eggs at week 12. The mean egg counts had increased to pre-treatment levels (Figure 2, panel C). The ERRs had decreased but were comparable, except for the PZQ plus As+SP group, which was significantly higher than the PZQ alone group, 55.8% vs 8.2%. See Table 3.

171 (39.4%) of the 434 patients cured by week six were re-infected by week 12. The reinfection risk was significantly higher in those who received PZQ plus DHAP than those who received PZQ alone (RR=1.50, 95%CI 1.03 to 2.19). See Table 3.

We did not observe any serious adverse events. 138 (25.6%) of the 540 children reported adverse events. A significantly higher proportion of those who received combination therapy reported an adverse event than those who received PZQ alone (see Table 3). Overall, 197 adverse events were reported. The majority of the adverse events were mild (81.2%) or moderate (18.8%), with the most common being abdominal pain (20.7%), headache (7.6%), and vomiting (3.1%). All adverse events did not require any treatment. Table 4 provides a summary of the reported adverse events by treatment group.

**Table 4.**
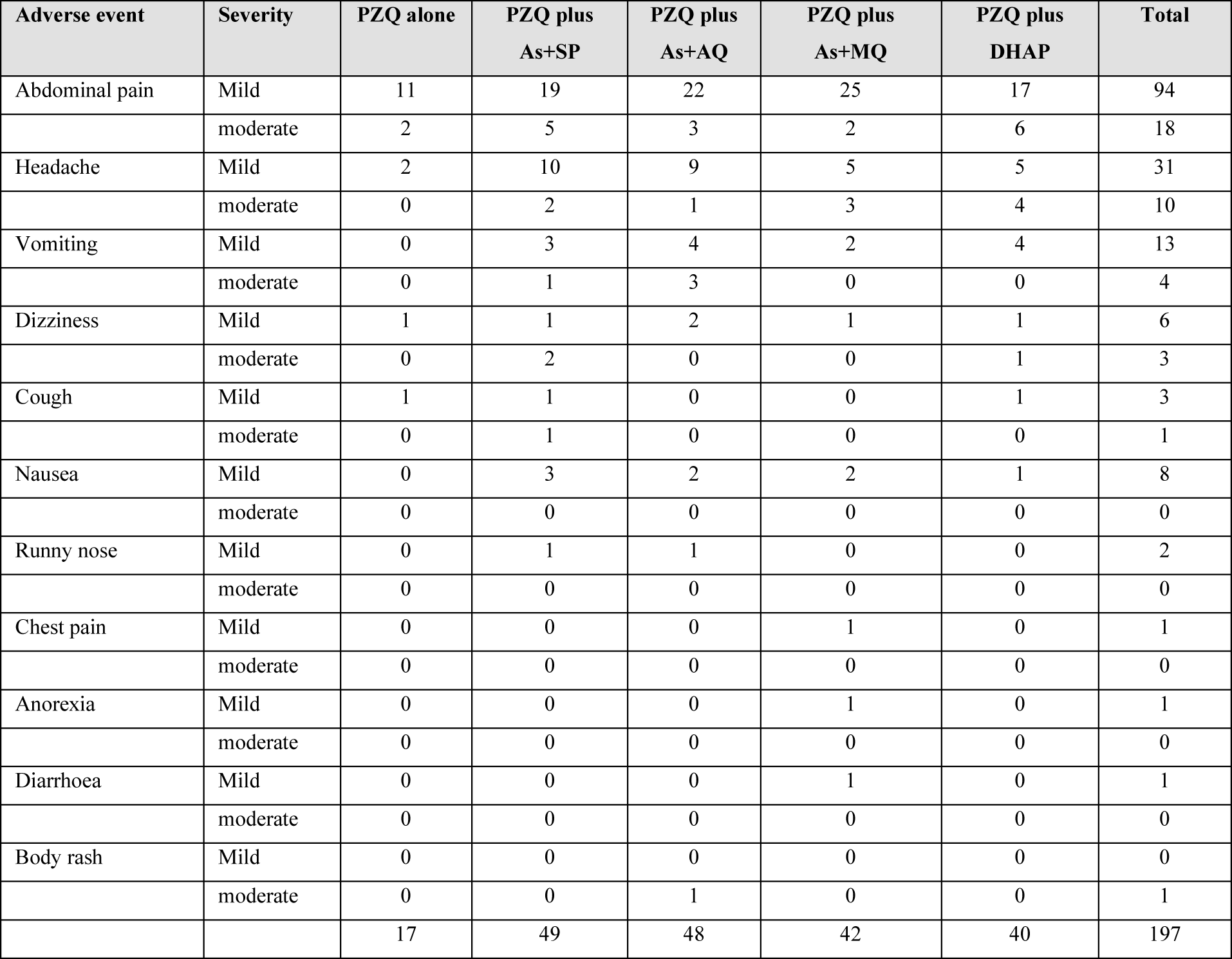
Summary of adverse events reported during the study. PZQ=praziquantel; As+SP=artesunate plus sulfalene/pyrimethamine; As+AQ= artesunate plus amodiaquine; As+MQ=artesunate plus mefloquine; DHAP=Dihydroartemisinin-piperaquine

| Adverse event | Severity | PZQ alone | PZQ plus<br>As+SP | PZQ plus<br>As+AQ | PZQ plus<br>As+MQ | PZQ plus<br>DHAP | Total |
| --- | --- | --- | --- | --- | --- | --- | --- |
| Abdominal pain | Mild | 11 | 19 | 22 | 25 | 17 | 94 |
|  | moderate | 2 | 5 | 3 | 2 | 6 | 18 |
| Headache | Mild | 2 | 10 | 9 | 5 | 5 | 31 |
|  | moderate | 0 | 2 | 1 | 3 | 4 | 10 |
| Vomiting | Mild | 0 | 3 | 4 | 2 | 4 | 13 |
|  | moderate | 0 | 1 | 3 | 0 | 0 | 4 |
| Dizziness | Mild | 1 | 1 | 2 | 1 | 1 | 6 |
|  | moderate | 0 | 2 | 0 | 0 | 1 | 3 |
| Cough | Mild | 1 | 1 | 0 | 0 | 1 | 3 |
|  | moderate | 0 | 1 | 0 | 0 | 0 | 1 |
| Nausea | Mild | 0 | 3 | 2 | 2 | 1 | 8 |
|  | moderate | 0 | 0 | 0 | 0 | 0 | 0 |
| Runny nose | Mild | 0 | 1 | 1 | 0 | 0 | 2 |
|  | moderate | 0 | 0 | 0 | 0 | 0 | 0 |
| Chest pain | Mild | 0 | 0 | 0 | 1 | 0 | 1 |
|  | moderate | 0 | 0 | 0 | 0 | 0 | 0 |
| Anorexia | Mild | 0 | 0 | 0 | 1 | 0 | 1 |
|  | moderate | 0 | 0 | 0 | 0 | 0 | 0 |
| Diarrhoea | Mild | 0 | 0 | 0 | 1 | 0 | 1 |
|  | moderate | 0 | 0 | 0 | 0 | 0 | 0 |
| Body rash | Mild | 0 | 0 | 0 | 0 | 0 | 0 |
|  | moderate | 0 | 0 | 1 | 0 | 0 | 1 |
|  |  | 17 | 49 | 48 | 42 | 40 | 197 |

## DISCUSSION

This is the first comparative head-to-head study of the efficacy and safety of praziquantel and four different ACTs in treating *S. mansoni* infection. Comparable cure rates but significantly increased ERRs were found between combination therapies and praziquantel at week 6. Noninferiority was declared for PZQ + As+MQ and PZQ + DHAP but not for PZQ plus As+SP or PZQ plus As+AQ. Study drugs were tolerated, and no serious adverse events were reported but children on combined treatment had significantly higher rates of adverse events than those on praziquantel monotherapy. Cure and egg reduction rates were not sustained through week 12.

The study found that single oral praziquantel therapy is effective and safe in treating intestinal schistosomiasis, with a cure rate of 82.5 %, and an ERR of 84.2 % six weeks post-treatment, consistent with previous studies [19]. Reinfection rates at week 12 were consistent with prior studies [20]. The treatment was most tolerable, with most patients reporting abdominal pain [19, 21].

Combination therapy administered over 3 days reduced infection intensity but not prevalence. Cure rates at 6 weeks were comparable, but ERRs were higher in children receiving combination treatment. Praziquantel combined with As+MQ or DHAP was non-inferior, suggesting they are potential alternatives to praziquantel monotherapy with ancillary benefits against schistosomulae. Our results contrast a recent study in which PZQ + DHAP had superior efficacy to PZQ alone in children with *S. mansoni* infections at 3- and 8 weeks post-treatment [8]. Adding dihydroartemisinin may have improved the bioavailability of PZQ [22]. However, it is unclear why children who received PZQ+DHAP in our study had an increased risk of reinfection at week 12.

Our study is the first to assess PZQ plus As+MQ for *S. mansoni.* We found relatively higher but comparable cure rates (88.7% vs 82.5%) with PZQ plus As+MQ at six weeks of follow-up. We anticipated PZQ plus As+MQ to show the most potent anti-schistosomal activity, as all three components have demonstrated effective anti-schistosomal properties in animal and human studies. For unclear reasons, our results contrast a study in Cote d’Ivoire where relatively low cure rates (33% vs 19%) were observed at 78-79 days (11 weeks) after treatment of children with *S. haematobium* using PZQ plus As+MQ compared to PZQ [9].

It is unclear why praziquantel plus As+SP and praziquantel plus As+AQ were inferior to PZQ alone in the treatment of *S.mansoni* infection in our study setting. The observed inferiority could be explained by the failure of these regimens to impact the intensity of infection. The mean difference in egg counts at week 6 in those who received PZQ combined with either As+SP or As+AQ were not different from PZQ and included both an increase as well as a decrease in egg counts.

The study found a higher proportion of adverse events in children on combination treatment for schistosomiasis, indicating the need for caution and safety monitoring in future ACT studies. The addition of drugs to praziquantel may have increased the frequency of adverse events but it is unclear if this would be lower if the study drugs were administered sequentially rather than simultaneously. The combination treatment was administered for three days, with more adverse events expected due to longer treatment duration, and drug-drug interactions. In previous studies, fewer adverse events were observed in the ACT group than in the PZQ group [10–12,14].

The study had limitations, including light-intensity infections in most children, an open-label design, single stool samples for diagnosis, potential misclassification of some children due to limitations of the Kato-Katz method, inability to measure treatment effect on juvenile worms, and short follow-up for long-term safety and efficacy outcomes. Inevitably, combination therapy will be costly and may be difficult to implement as part of mass drug administration. For this study, single dose PZQ and a 3-day course of ACT were co-administered, and this may have impacted compliance and influenced the adverse events reporting.

The current schistosomiasis control strategy relies on praziquantel for preventive chemotherapy. Combination therapy using praziquantel with artesunate plus mefloquine or praziquantel with dihydroartemisinin-piperaquine is a promising complementary transmission control strategy but further research is needed to investigate strategies to improve the effectiveness and safety outcomes. The study raises concerns about the safety, effectiveness, and interactions of drugs used in combination therapy for intestinal schistosomiasis. Further research is needed to validate findings across schistosome species, determine dosing intervals, explore the potential for antimalarial drug resistance, and evaluate the effectiveness of sequential, simultaneous, or single-dose administration. Combination therapy could be beneficial in malaria-free areas, in patients with both malaria and schistosomiasis, as a backup treatment for PZQ failure, for travellers to schistosomiasis-prone regions, and for seasonal chemotherapy in areas where malaria and schistosomiasis coexist. Combination therapy comprising of PZQ and an ACT is likely to transform the schistosomiasis control strategy from morbidity to transmission control and may accelerate the global schistosomiasis elimination agenda. In conclusion, we found that praziquantel with artesunate plus mefloquine and praziquantel with dihydroartemisinin-piperaquine were non-inferior to praziquantel but were associated with a risk of adverse events. Overall, the role of ACTs in schistosomiasis control remains unclear.

## Data Availability

The datasets used and analyzed during the current study are available from the corresponding author upon reasonable request.

## Ethical statement

The trial is registered with the Pan-African Clinical Trials Registry, PACTR202001919442161. Ethics approval was obtained from the Scientific and Ethics Review Unit of the Kenya Medical Research Institute (SERU #3507). The trial was conducted per the Declaration of Helsinki and ICH Good Clinical Practice. The Kirinyaga County Commissioner, the Kirinyaga County Directorates of Health and Education, and the respective school administration granted permission to conduct the study. Written informed consent was obtained from the parents/guardians, and verbal assent from all eligible children.

## Author Contributions

CO initiated the idea, wrote the study protocol, and sourced for funding. CO, EM, PW, and VW supervised the data collection. CO and VW analyzed and interpreted the data. CO drafted the manuscript. All authors contributed to the writing of the paper and approved the final version.

## Acknowledgments

We thank the children and schools that participated in this study. This study is published with the permission of the Director-General of KEMRI.

## Consent for publication

Not applicable

